# Olfactory Dysfunction in Primary Ciliary Dyskinesia: A Systematic Review and Meta-analysis

**DOI:** 10.64898/2026.08.17.26355624

**Authors:** Arshad Zubair, Katherine Whitcroft, Eishaan Bhargava, Grace Khong

## Abstract

**Objective:** Primary ciliary dyskinesia (PCD) is a rare congenital motile ciliopathy associated with significant sinopulmonary disease, alongside marked genetic and phenotypic variability. Olfactory dysfunction in PCD remains poorly characterised. This systematic review aimed to synthesise the available evidence on the prevalence, assessment methods, and clinical correlates of olfactory dysfunction in individuals with PCD.

**Methods:** A PRISMA-compliant systematic review was conducted. Medline, Embase, Scopus, Web of Science, and the Cochrane Library were searched up to 28/02/26. Observational studies and clinical trials reporting on olfactory function in confirmed PCD were included. Data were extracted, and study quality was assessed using the Newcastle-Ottawa Scale (NOS). A random-effects meta-analysis using the Freeman-Tukey double arcsine transformation was performed to calculate the pooled prevalence, 95% confidence interval (CI), and 95% prediction interval (PI). The review protocol was registered with PROSPERO (CRID: 1006332).

**Results:** Of 159 records screened, 12 studies (n=865) met the inclusion criteria. The prevalence of olfactory dysfunction varied widely, from 1.5% to 100%. The overall pooled prevalence was 43.4% (95% CI 25.2%–62.5%), but with a very wide 95% prediction interval of 0.1%–99.0%, reflecting significant heterogeneity (I²=96.2%). Objective psychophysical tests (n=6 studies) identified a pooled prevalence of 66.1% (95% CI 55.5%–76.0%; 95% PI 38.4%–88.9%), which was significantly higher than that from patient-reported outcome measures (30.5%; 95% CI 11.4%–54.0%; 95% PI 11.4%–97.1%) and clinical history (5.6%; 95% CI 0.2%–24.8%). Older age, specific ciliary ultrastructural defects, and greater sinonasal disease burden on imaging were associated with worse olfactory function.

**Conclusion:** Olfactory dysfunction is a highly prevalent yet profoundly under-recognised comorbidity in Primary Ciliary Dyskinesia, with over two-thirds of patients affected when assessed objectively. The pathophysiology extends beyond simple conductive obstruction to likely include a primary neurosensory deficit linked to the underlying genetic defect. These findings carry a clear mandate for clinicians to integrate objective olfactory testing into routine PCD care to improve patient safety and quality of life.

## Introduction

Primary ciliary dyskinesia (PCD) is a rare, genetically heterogeneous congenital disorder characterised by impaired ciliary structure and function ^(1)^. With an estimated prevalence of 1 in 15,000 to 30,000 live births, this predominantly autosomal recessive condition leads to defective mucociliary clearance throughout the respiratory tract ^(2)^. Clinical manifestations typically begin in early infancy and include chronic wet cough, neonatal respiratory distress, recurrent otitis media, and the development of bronchiectasis. Over 75% of all individuals with PCD also suffer from chronic rhinosinusitis (CRS), which contributes significantly to the disease burden ^(3)^.

Olfactory dysfunction is a well-established and common symptom of CRS, affecting patient safety and quality of life ^(4)^. However, the sense of smell in the context of PCD has historically been under-investigated. The olfactory neuroepithelium, located in the superior nasal cavity, contains cilia on its olfactory sensory neurons. While these are non-motile primary cilia, distinct from the motile respiratory cilia affected in PCD, the surrounding environment of chronic inflammation, viscous mucus, and potential oedema secondary to CRS can create a significant conductive barrier to odourant molecules reaching the neuroepithelium ^(5)^.

More recent evidence suggests that the pathophysiology of olfactory loss in PCD may be more complex than conductive loss alone. Associations between the severity of olfactory loss and specific ciliary ultrastructural defects on transmission electron microscopy (TEM) have been reported, raising the possibility of a primary neurosensory component to the olfactory deficit ^(6)^. Studies using objective psychophysical testing have revealed a high prevalence of olfactory dysfunction in PCD, with rates approaching or exceeding 70% in several cohorts, which is disproportionately high compared to patient-reported symptoms^(6,7)^.

Given the emerging evidence of a high, under-recognised prevalence and a potentially complex pathophysiology, a comprehensive evaluation of literature is warranted. This systematic review aims to synthesise all available evidence on the prevalence of olfactory dysfunction in individuals with PCD, evaluate the different assessment methods used, and identify the clinical, radiological, and genetic correlates of olfactory impairment in this population.

## Methods

This systematic review was conducted and reported in accordance with the Preferred Reporting Items for Systematic Reviews and Meta-Analyses (PRISMA) 2020 statement ^(8)^. The review protocol was registered prospectively with the International Prospective Register of Systematic Reviews (PROSPERO; CRD420251006332)^(9)^.

### Eligibility Criteria

Studies were included if they met the following criteria based on the Population, Exposure, Comparator, and Outcome (PECO) framework:

Population: Individuals of any age with a confirmed diagnosis of PCD, based on established international guidelines (e.g., genetic testing, transmission electron microscopy, or nasal nitric oxide measurements)^(2)^.

Exposure: Impaired motile ciliary structure and function, as a consequence of underlying genetic defect in confirmed PCD.

Comparator: Non-PCD healthy controls, if reported. Outcome:

### Primary: Olfactory function measured via validated tools

Secondary: Correlations with sinonasal symptoms, quality of life metrics, or genetic variants.

We included observational study designs (cross-sectional, cohort, case-control) and clinical trials reporting olfactory function. Studies not available in English, case reports, and review articles or conference abstracts without primary data were excluded. No minimum participant threshold was applied.

### Search Strategy

A comprehensive search of multiple electronic databases was performed from their inception to 28/2/26. The databases included Medline, Embase, Scopus, Web of Science, and the Cochrane Library. To identify unpublished or ongoing trials, grey literature sources including ClinicalTrials.gov and OpenGrey were also searched. The search strategy combined Medical Subject Headings (MeSH) and Emtree terms with free-text keywords. The core search logic was: (“Primary Ciliary Dyskinesia” OR “Kartagener Syndrome”) AND (“Olfaction” OR “Smell” OR “Anosmia”), adapted for each database’s specific syntax. The reference lists of all included articles and relevant reviews were also manually screened to identify additional studies.

### Study Selection and Data Extraction

All identified records were imported into a reference management software (Rayyan) for duplicate removal and screening. Three reviewers (A.Z, E.B, G.K.) independently screened the titles and abstracts of the remaining records against the eligibility criteria. The full texts of potentially relevant articles were then retrieved and assessed for final inclusion by the same three reviewers. Any disagreements at either stage were resolved through discussion or, if necessary, by a fourth reviewer (K.W.).

A standardised data extraction form was developed in Microsoft Excel. One reviewer (A.Z.) extracted the following data from each study: first author and year of publication, study design, sample size, participant demographics (age, sex), PCD diagnostic criteria, olfactory assessment method, prevalence or rate of olfactory dysfunction, comparator group details, sinonasal assessment data (e.g., SNOT-22, Lund-Mackay scores), and any reported associations with genotype or other clinical parameters.

The methodological quality and risk of bias of the included studies were independently assessed by two reviewers (A.Z., E.B.) using the Newcastle-Ottawa Scale (NOS) adapted for cross-sectional and cohort studies ^(10)^. The scale evaluates three domains: (1) Selection of the study groups (representativeness, ascertainment of exposure; maximum 4 stars); (2) Comparability of the groups (control for confounders; maximum 2 stars); and (3) Ascertainment of the Outcome (assessment method, follow-up; maximum 3 stars). Studies were categorised as having a low (score ≥7), moderate (score 5–6), or high (score ≤4) risk of bias. Discrepancies were resolved by consensus.

### Data Synthesis and Statistical Analysis

Data were synthesised narratively and quantitatively. A random-effects meta-analysis was performed to calculate a pooled prevalence of olfactory dysfunction. The analysis was conducted using the Freeman-Tukey double arcsine transformation to stabilise variance. In addition to the 95% confidence interval (CI) for the pooled estimate, a 95% prediction interval (PI) was calculated to estimate the range of true prevalence in future studies. Heterogeneity between studies was assessed using the I² statistic, with values of <25%, 25%–75%, and >75% considered low, moderate, and high heterogeneity, respectively. Pre-specified subgroup analyses were conducted based on the method of olfactory assessment: (1) objective psychophysical testing (e.g., UPSIT, Sniffin’ Sticks), (2) validated PROMs (e.g., SNOT-22, FOLLOW-PCD), and (3) clinical history. All statistical analyses were performed using R (version 4.5.3; R Foundation for Statistical Computing, Vienna, Austria). A leave-one-out sensitivity analysis was also performed. The certainty of evidence was independently assessed using the Grading of Recommendations Assessment, Development and Evaluation (GRADE) framework, rating evidence as high, moderate, low, or very low based on five domains: risk of bias, inconsistency, indirectness, imprecision, and publication bias.

## Results

### Study Selection and Characteristics

The search identified 159 records. After screening, 12 studies were included, comprising 865 participants with PCD. The PRISMA flow diagram is shown in Figure 1. Study designs included cross-sectional (n=6), prospective cohorts (n=3), and case series (n=3). Six studies used objective psychophysical tests, four used PROMs, and two relied on clinical history. Study characteristics are detailed in Table 1.

**Figure 1:**
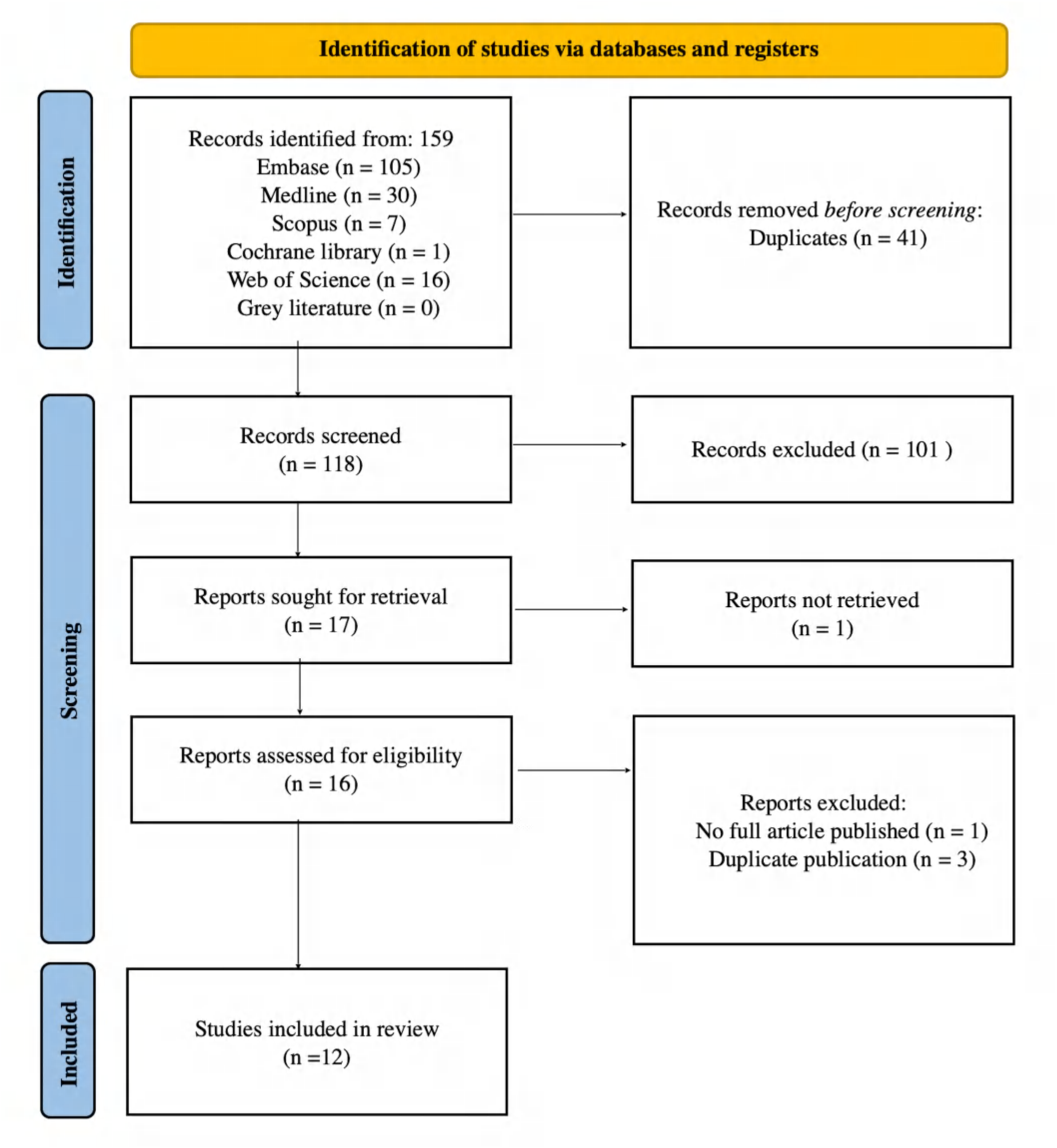
PRISMA 2020 flow diagram of the study selection process.

**Table 1:**
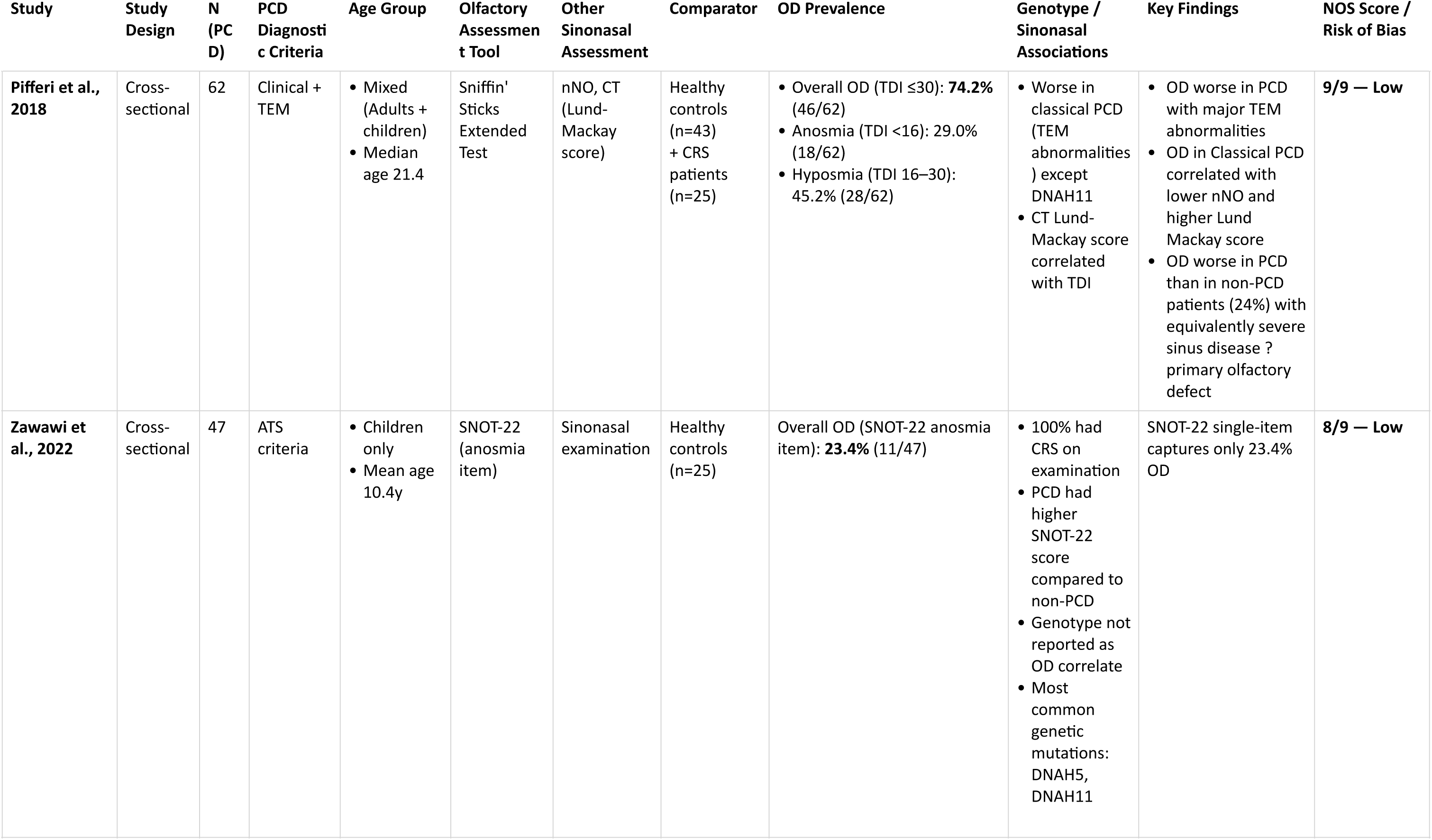
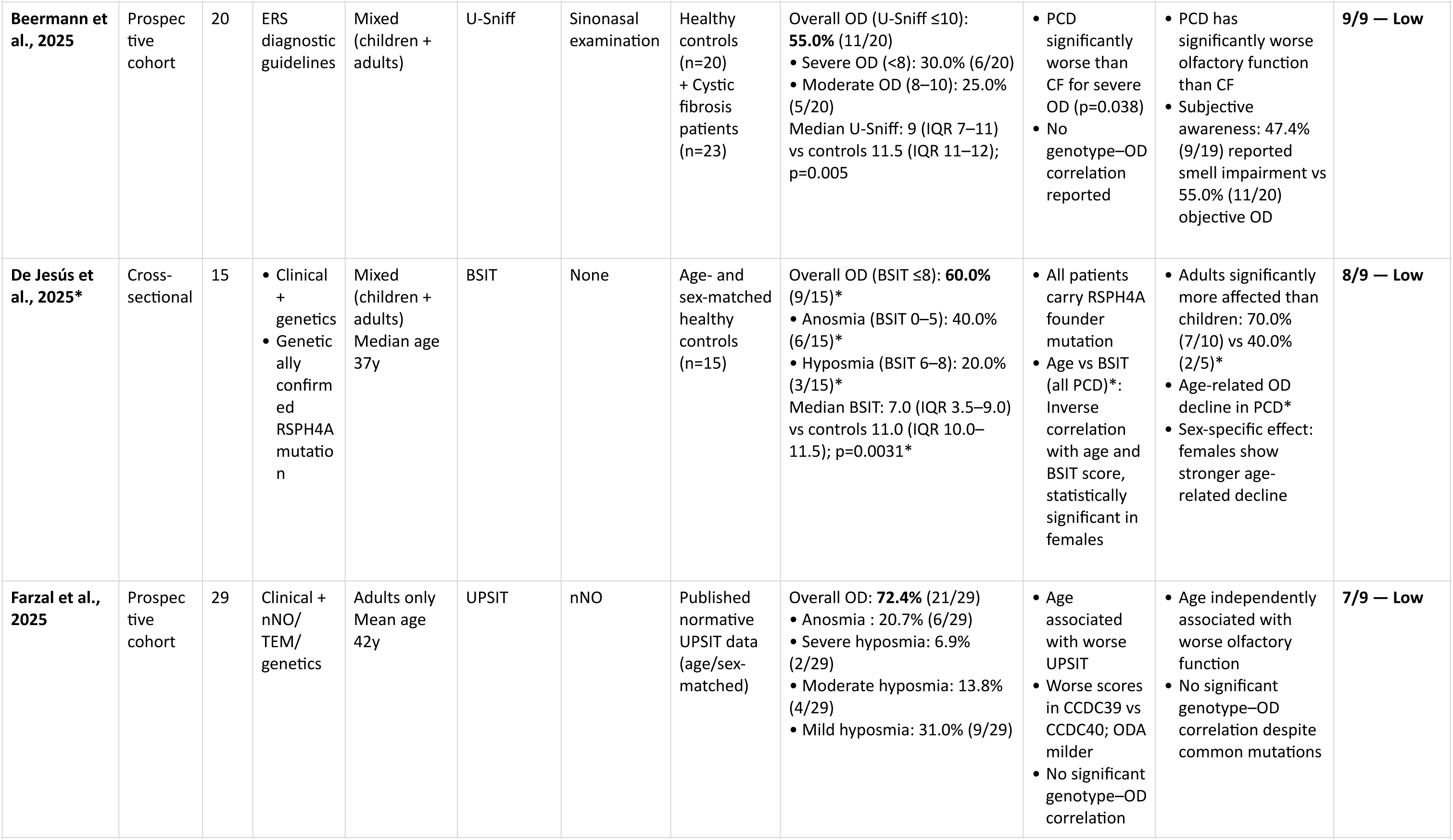
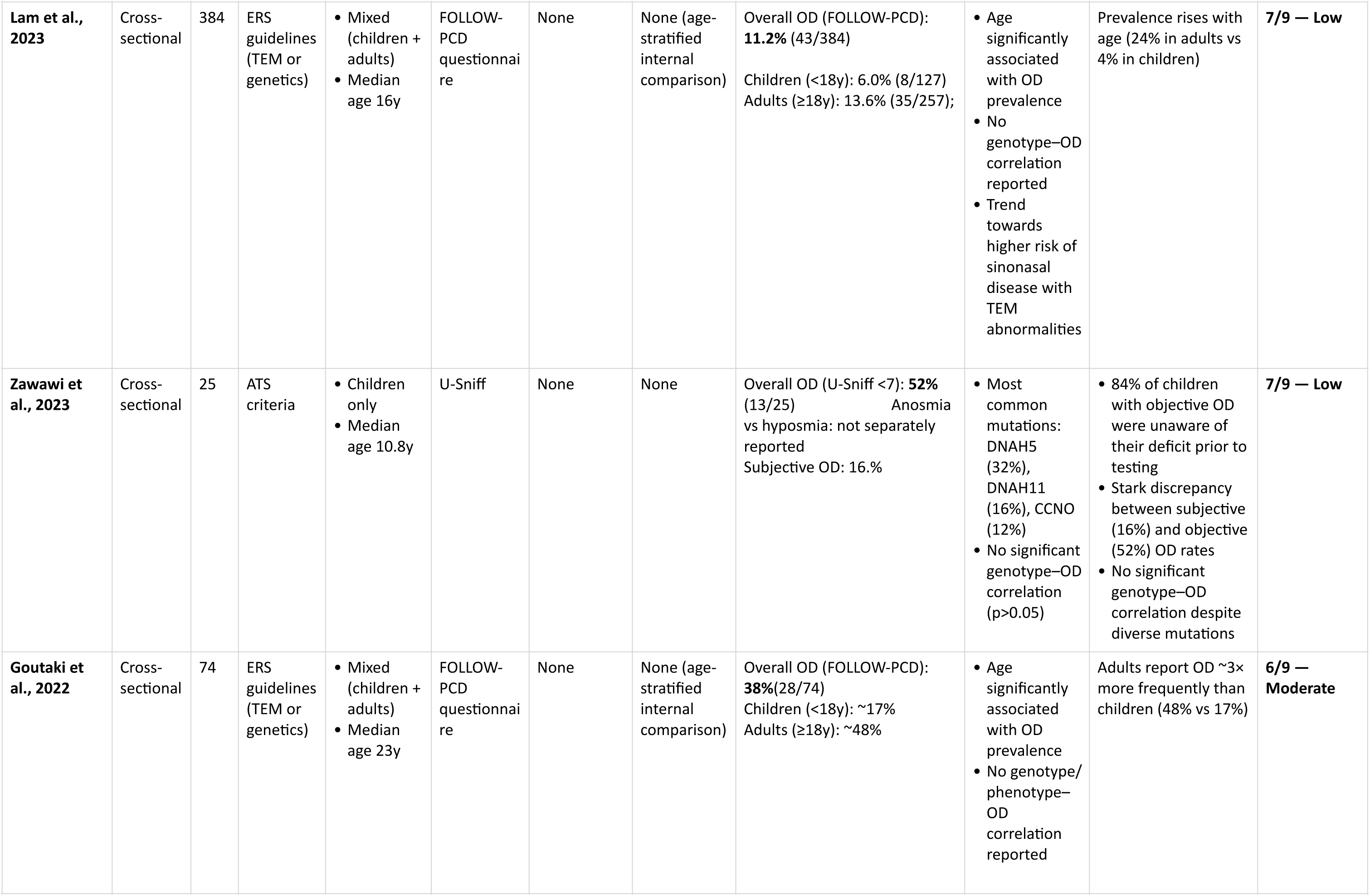
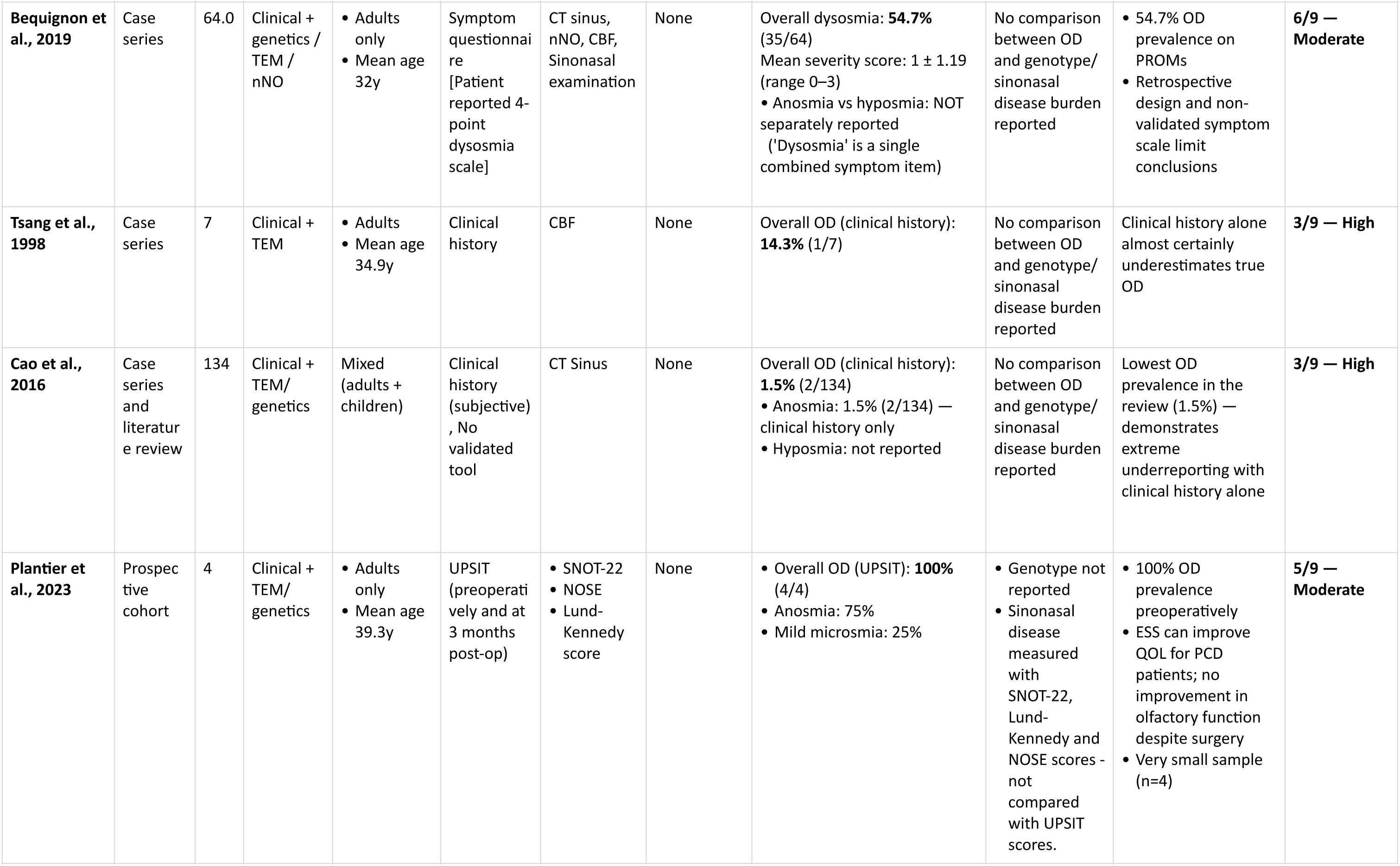
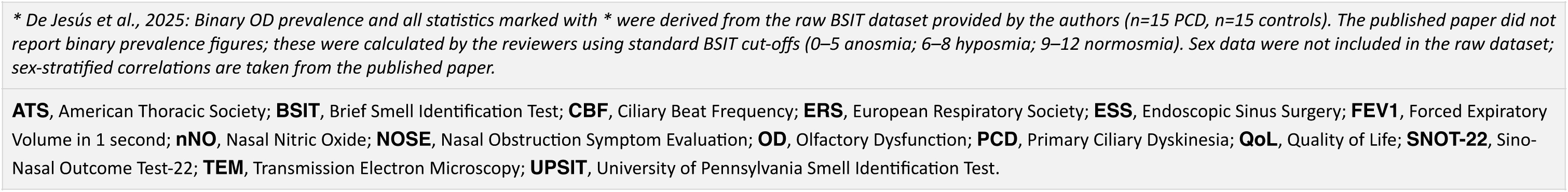
Characteristics of Included Studies (Total of 12 Studies)

| Study | Study Design | N (PCD) | PCD Diagnostic Criteria | Age Group | Olfactory Assessment Tool | Other Sinonasal Assessment | Comparator | OD Prevalence | Genotype / Sinonasal Associations | Key Findings | NOS Score / Risk of Bias |
| --- | --- | --- | --- | --- | --- | --- | --- | --- | --- | --- | --- |
| Pifferi et al., 2018 | Cross-sectional | 62 | Clinical + TEM | Mixed (Adults + children)<br>Median age 21.4 | Sniffin' Sticks Extended Test | nNO, CT (Lund-Mackay score) | Healthy controls (n=43) + CRS patients (n=25) | Overall OD (TDI $\leq 30$): **74.2%** (46/62)<br>Anosmia (TDI $< 16$): 29.0% (18/62)<br>Hyposmia (TDI 16–30): 45.2% (28/62) | Worse in classical PCD (TEM abnormalities) except DNAH11<br>CT Lund-Mackay score correlated with TDI | OD worse in PCD with major TEM abnormalities<br>OD in Classical PCD correlated with lower nNO and higher Lund-Mackay score<br>OD worse in PCD than in non-PCD patients (24%) with equivalently severe sinus disease ? primary olfactory defect | **9/9 — Low** |
| Zawawi et al., 2022 | Cross-sectional | 47 | ATS criteria | Children only<br>Mean age 10.4y | SNOT-22 (anosmia item) | Sinonasal examination | Healthy controls (n=25) | Overall OD (SNOT-22 anosmia item): **23.4%** (11/47) | 100% had CRS on examination<br>PCD had higher SNOT-22 score compared to non-PCD<br>Genotype not reported as OD correlate<br>Most common genetic mutations: DNAH5, DNAH11 | SNOT-22 single-item captures only 23.4% OD | **8/9 — Low** |
| **Beermann et al., 2025** | Prospective cohort | 20 | ERS diagnostic guidelines | Mixed (children + adults) | U-Sniff | Sinonasal examination | Healthy controls (n=20) + Cystic fibrosis patients (n=23) | Overall OD (U-Sniff ≤10): **55.0%** (11/20)<br>• Severe OD (<8): 30.0% (6/20)<br>• Moderate OD (8–10): 25.0% (5/20)<br>Median U-Sniff: 9 (IQR 7–11) vs controls 11.5 (IQR 11–12); p=0.005 | • PCD significantly worse than CF for severe OD (p=0.038)<br>• No genotype–OD correlation reported | • PCD has significantly worse olfactory function than CF<br>• Subjective awareness: 47.4% (9/19) reported smell impairment vs 55.0% (11/20) objective OD | **9/9 — Low** |
| **De Jesús et al., 2025\*** | Cross-sectional | 15 | • Clinical + genetics<br>• Genetically confirmed RSPH4A mutation | Mixed (children + adults)<br>Median age 37y | BSIT | None | Age- and sex-matched healthy controls (n=15) | Overall OD (BSIT ≤8): **60.0%** (9/15)*<br>• Anosmia (BSIT 0–5): 40.0% (6/15)*<br>• Hyposmia (BSIT 6–8): 20.0% (3/15)*<br>Median BSIT: 7.0 (IQR 3.5–9.0) vs controls 11.0 (IQR 10.0–11.5); p=0.0031* | • All patients carry RSPH4A founder mutation<br>• Age vs BSIT (all PCD)*: Inverse correlation with age and BSIT score, statistically significant in females | • Adults significantly more affected than children: 70.0% (7/10) vs 40.0% (2/5)*<br>• Age-related OD decline in PCD*<br>• Sex-specific effect: females show stronger age-related decline | **8/9 — Low** |
| **Farzal et al., 2025** | Prospective cohort | 29 | Clinical + nNO/TEM/genetics | Adults only<br>Mean age 42y | UPSIT | nNO | Published normative UPSIT data (age/sex-matched) | Overall OD: **72.4%** (21/29)<br>• Anosmia : 20.7% (6/29)<br>• Severe hyposmia: 6.9% (2/29)<br>• Moderate hyposmia: 13.8% (4/29)<br>• Mild hyposmia: 31.0% (9/29) | • Age associated with worse UPSIT<br>• Worse scores in CCDC39 vs CCDC40; ODA milder<br>• No significant genotype–OD correlation | • Age independently associated with worse olfactory function<br>• No significant genotype–OD correlation despite common mutations | **7/9 — Low** |
| **Lam et al., 2023** | Cross-sectional | 384 | ERS guidelines (TEM or genetics) | Mixed (children + adults)<br>Median age 16y | FOLLOW-PCD questionnaire | None | None (age-stratified internal comparison) | Overall OD (FOLLOW-PCD): **11.2%** (43/384)<br>Children (<18y): 6.0% (8/127)<br>Adults (≥18y): 13.6% (35/257); | Age significantly associated with OD prevalence<br>No genotype–OD correlation reported<br>Trend towards higher risk of sinonasal disease with TEM abnormalities | Prevalence rises with age (24% in adults vs 4% in children) | **7/9 — Low** |
| **Zawawi et al., 2023** | Cross-sectional | 25 | ATS criteria | Children only<br>Median age 10.8y | U-Sniff | None | None | Overall OD (U-Sniff <7): **52%** (13/25) Anosmia vs hyposmia: not separately reported<br>Subjective OD: 16.6% | Most common mutations: DNAH5 (32%), DNAH11 (16%), CCNO (12%)<br>No significant genotype–OD correlation (p>0.05) | 84% of children with objective OD were unaware of their deficit prior to testing<br>Stark discrepancy between subjective (16%) and objective (52%) OD rates<br>No significant genotype–OD correlation despite diverse mutations | **7/9 — Low** |
| **Goutaki et al., 2022** | Cross-sectional | 74 | ERS guidelines (TEM or genetics) | Mixed (children + adults)<br>Median age 23y | FOLLOW-PCD questionnaire | None | None (age-stratified internal comparison) | Overall OD (FOLLOW-PCD): **38%**(28/74)<br>Children (<18y): ~17%<br>Adults (≥18y): ~48% | Age significantly associated with OD prevalence<br>No genotype/phenotype–OD correlation reported | Adults report OD ~3× more frequently than children (48% vs 17%) | **6/9 — Moderate** |
| **Bequignon et al., 2019** | Case series | 64.0 | Clinical + genetics / TEM / nNO | Adults only<br>Mean age 32y | Symptom questionnaire [Patient reported 4-point dysosmia scale] | CT sinus, nNO, CBF, Sinonasal examination | None | Overall dysosmia: **54.7%** (35/64)<br>Mean severity score: $1 \pm 1.19$ (range 0–3)<br>Anosmia vs hyposmia: NOT separately reported ('Dysosmia' is a single combined symptom item) | No comparison between OD and genotype/sinonasal disease burden reported | 54.7% OD prevalence on PROMs<br>Retrospective design and non-validated symptom scale limit conclusions | **6/9 — Moderate** |
| **Tsang et al., 1998** | Case series | 7 | Clinical + TEM | Adults<br>Mean age 34.9y | Clinical history | CBF | None | Overall OD (clinical history): **14.3%** (1/7) | No comparison between OD and genotype/sinonasal disease burden reported | Clinical history alone almost certainly underestimates true OD | **3/9 — High** |
| **Cao et al., 2016** | Case series and literature review | 134 | Clinical + TEM/genetics | Mixed (adults + children) | Clinical history (subjective), No validated tool | CT Sinus | None | Overall OD (clinical history): **1.5%** (2/134)<br>Anosmia: 1.5% (2/134) — clinical history only<br>Hyposmia: not reported | No comparison between OD and genotype/sinonasal disease burden reported | Lowest OD prevalence in the review (1.5%) — demonstrates extreme underreporting with clinical history alone | **3/9 — High** |
| **Plantier et al., 2023** | Prospective cohort | 4 | Clinical + TEM/genetics | Adults only<br>Mean age 39.3y | UPSIT (preoperatively and at 3 months post-op) | SNOT-22<br>NOSE<br>Lund-Kennedy score | None | Overall OD (UPSIT): **100%** (4/4)<br>Anosmia: 75%<br>Mild microsmia: 25% | Genotype not reported<br>Sinonasal disease measured with SNOT-22, Lund-Kennedy and NOSE scores - not compared with UPSIT scores. | 100% OD prevalence preoperatively<br>ESS can improve QOL for PCD patients; no improvement in olfactory function despite surgery<br>Very small sample (n=4) | **5/9 — Moderate** |

*\* De Jesús et al., 2025: Binary OD prevalence and all statistics marked with \* were derived from the raw BSIT dataset provided by the authors (n=15 PCD, n=15 controls). The published paper did not report binary prevalence figures; these were calculated by the reviewers using standard BSIT cut-offs (0–5 anosmia; 6–8 hyposmia; 9–12 normosmia). Sex data were not included in the raw dataset; sex-stratified correlations are taken from the published paper.*
**ATS**, American Thoracic Society; **BSIT**, Brief Smell Identification Test; **CBF**, Ciliary Beat Frequency; **ERS**, European Respiratory Society; **ESS**, Endoscopic Sinus Surgery; **FEV1**, Forced Expiratory Volume in 1 second; **nNO**, Nasal Nitric Oxide; **NOSE**, Nasal Obstruction Symptom Evaluation; **OD**, Olfactory Dysfunction; **PCD**, Primary Ciliary Dyskinesia; **QoL**, Quality of Life; **SNOT-22**, Sino-Nasal Outcome Test-22; **TEM**, Transmission Electron Microscopy; **UPSIT**, University of Pennsylvania Smell Identification Test.

### Risk of Bias Assessment

Using the NOS, seven studies (58%) were rated as having a low risk of bias, three (25%) as moderate, and two (17%) as high (Figure 2). Common sources of potential bias were the lack of a comparator group and the use of subjective, non-validated methods for assessing olfactory function.

**Figure 2:**
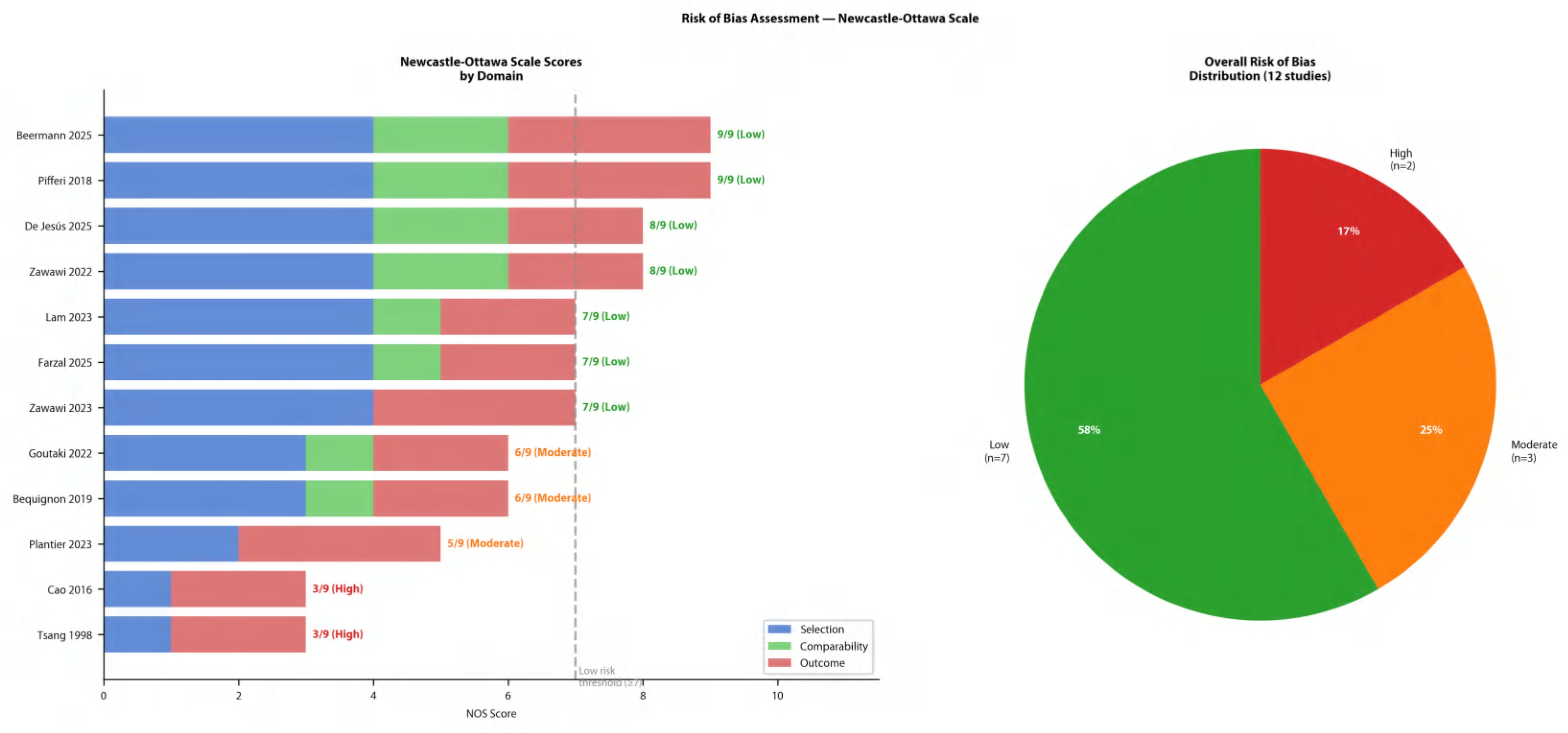
Risk of bias assessment of included studies using the Newcastle-Ottawa Scale (NOS).

A funnel plot was constructed to assess the potential for publication bias (Figure 3).The funnel plot demonstrated mild asymmetry, with a cluster of smaller studies (higher standard error) distributed towards the left of the pooled estimate, and one outlying study (Plantier, 2023; n=4) falling outside the pseudo-confidence funnel on the right (Figure 3). The observed asymmetry most likely reflects the substantial true heterogeneity (I²=96.2%) and differences in study population and assessment method rather than selective publication bias.

**Figure 3:**
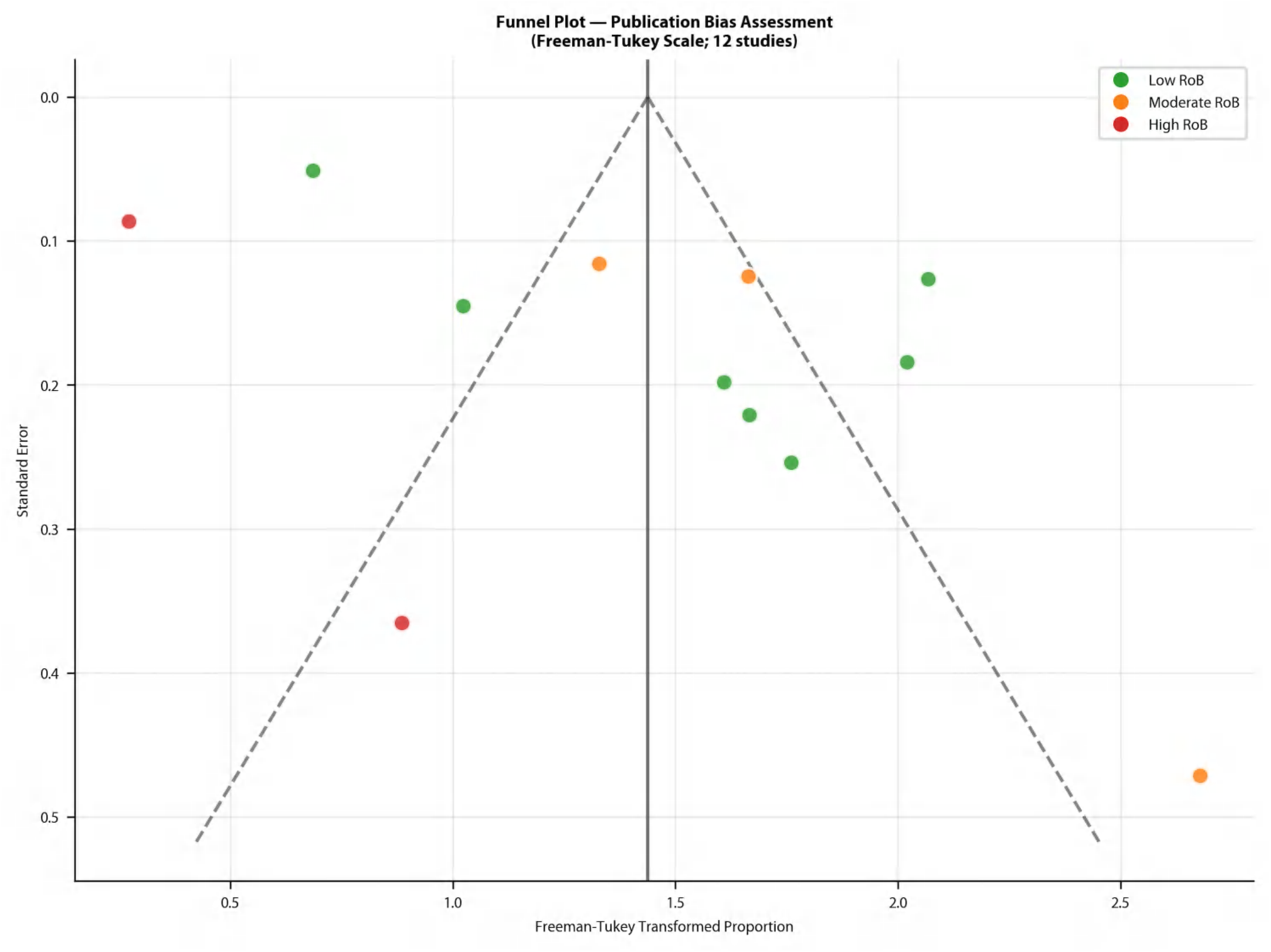
Funnel plot demonstrating publication bias in the included studies

### Prevalence of Olfactory Dysfunction

The prevalence of olfactory dysfunction in individual studies varied from 1.5% to 100%. The random-effects meta-analysis yielded an overall pooled prevalence of 43.4% (95% CI 25.2%–62.5%). However, the 95% prediction interval was extremely wide at 0.1% to 99.0%, reflecting the substantial heterogeneity between studies (I² = 96.2%, p < 0.001) (Figure 4).

**Figure 4:**
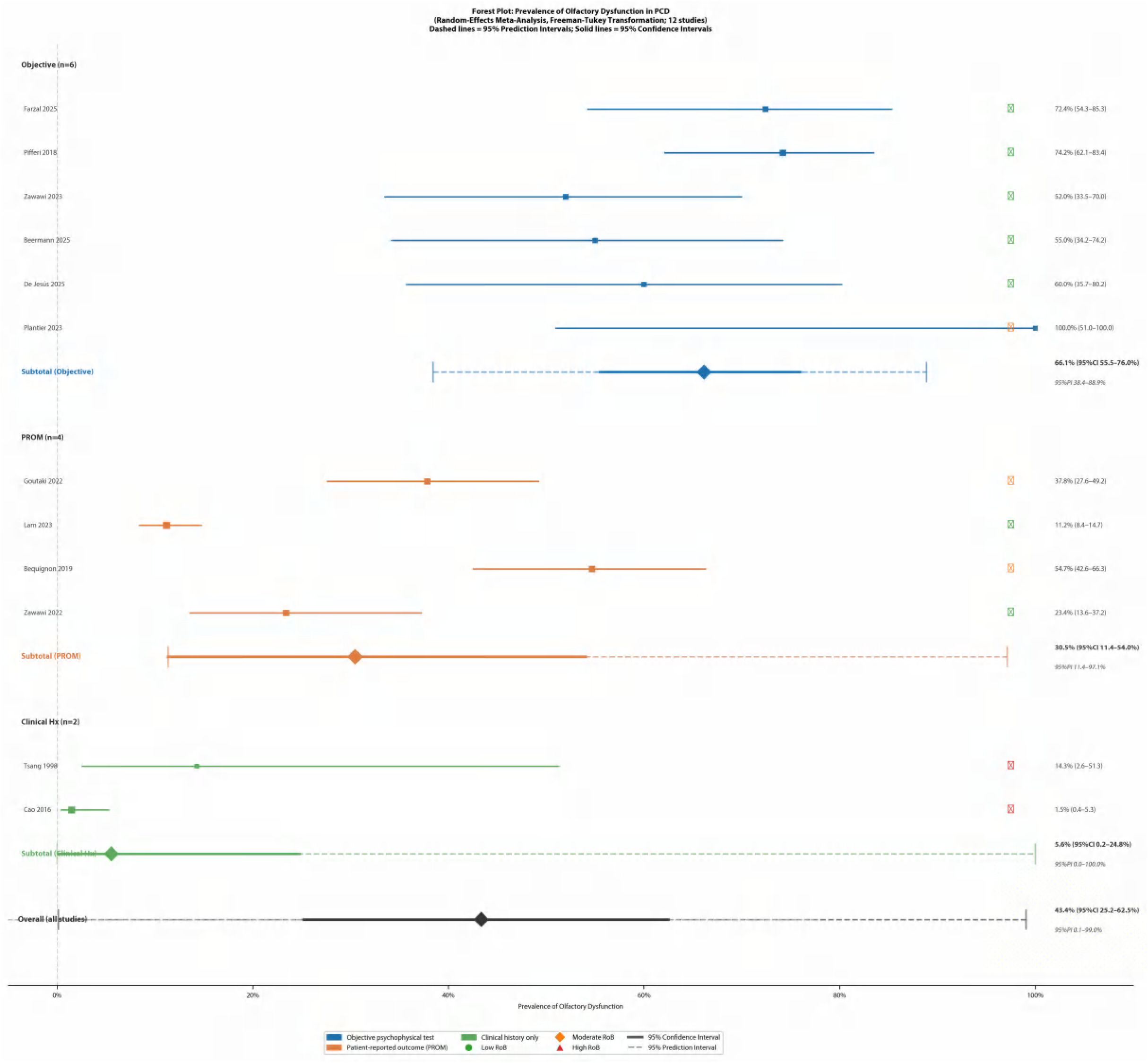
Forest plot of the prevalence of olfactory dysfunction in PCD. The plot shows individual study prevalence (squares), the pooled 95% confidence intervals (solid diamonds), and the 95% prediction intervals (dashed lines).

### Subgroup and Sensitivity Analysis

Subgroup analysis revealed significant differences by assessment method (Figure 5). Objective testing yielded the highest pooled prevalence at 66.1% (95% CI 55.5%–76.0%; 95% PI 38.4%–88.9%; I²=40.5%). PROMs yielded a prevalence of 30.5% (95% CI 11.4%–54.0%; 95% PI 11.4%–97.1%; I²=95.7%), and clinical history yielded 5.6% (95% CI 0.2%–24.8%; I²=62.6%). A leave-one-out sensitivity analysis confirmed the overall estimate was robust and not driven by any single study (Figure 6).

**Figure 5:**
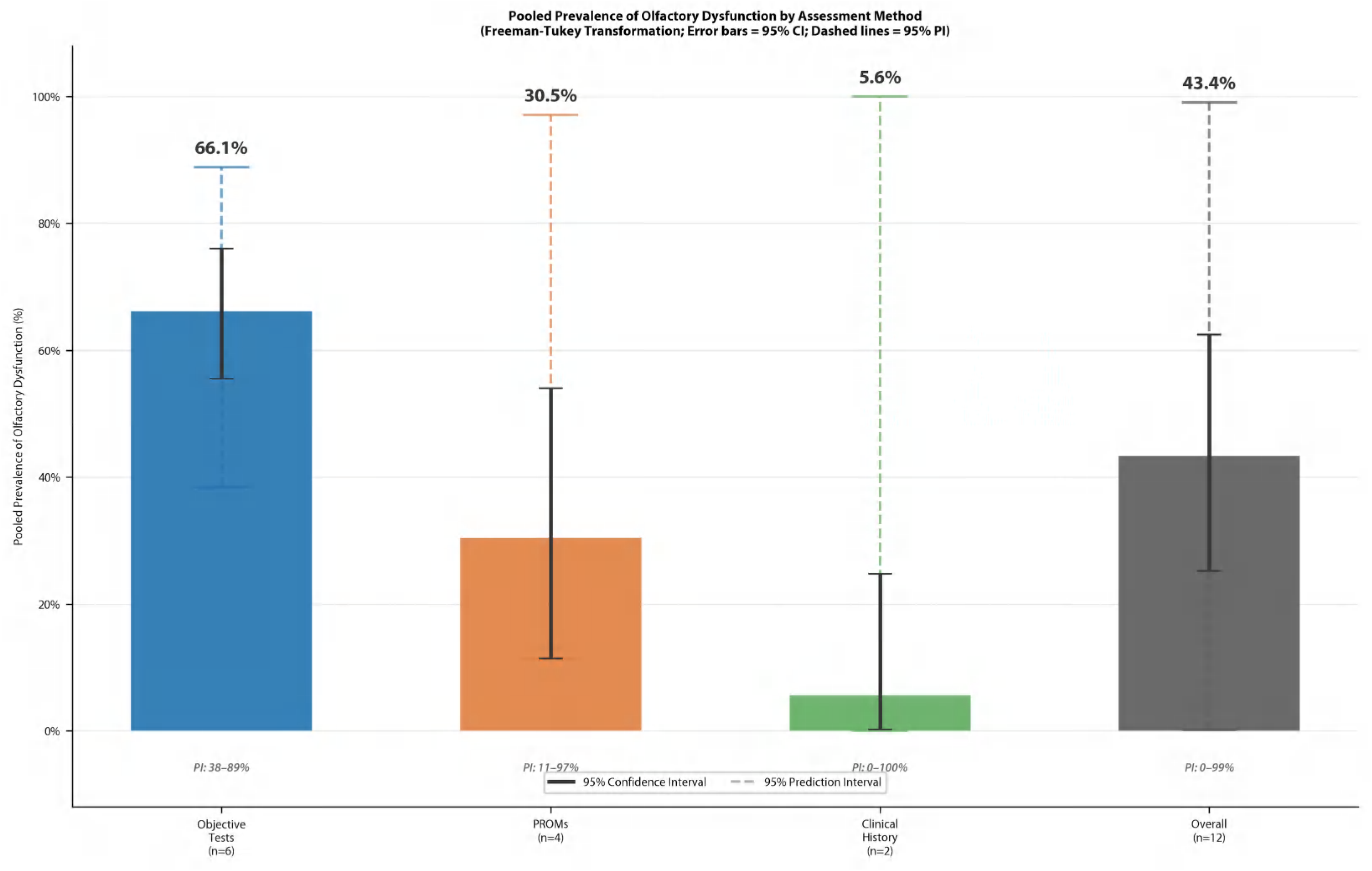
Pooled prevalence of olfactory dysfunction by assessment method. Error bars represent the 95% CI; dashed lines represent the 95% PI.

**Figure 6:**
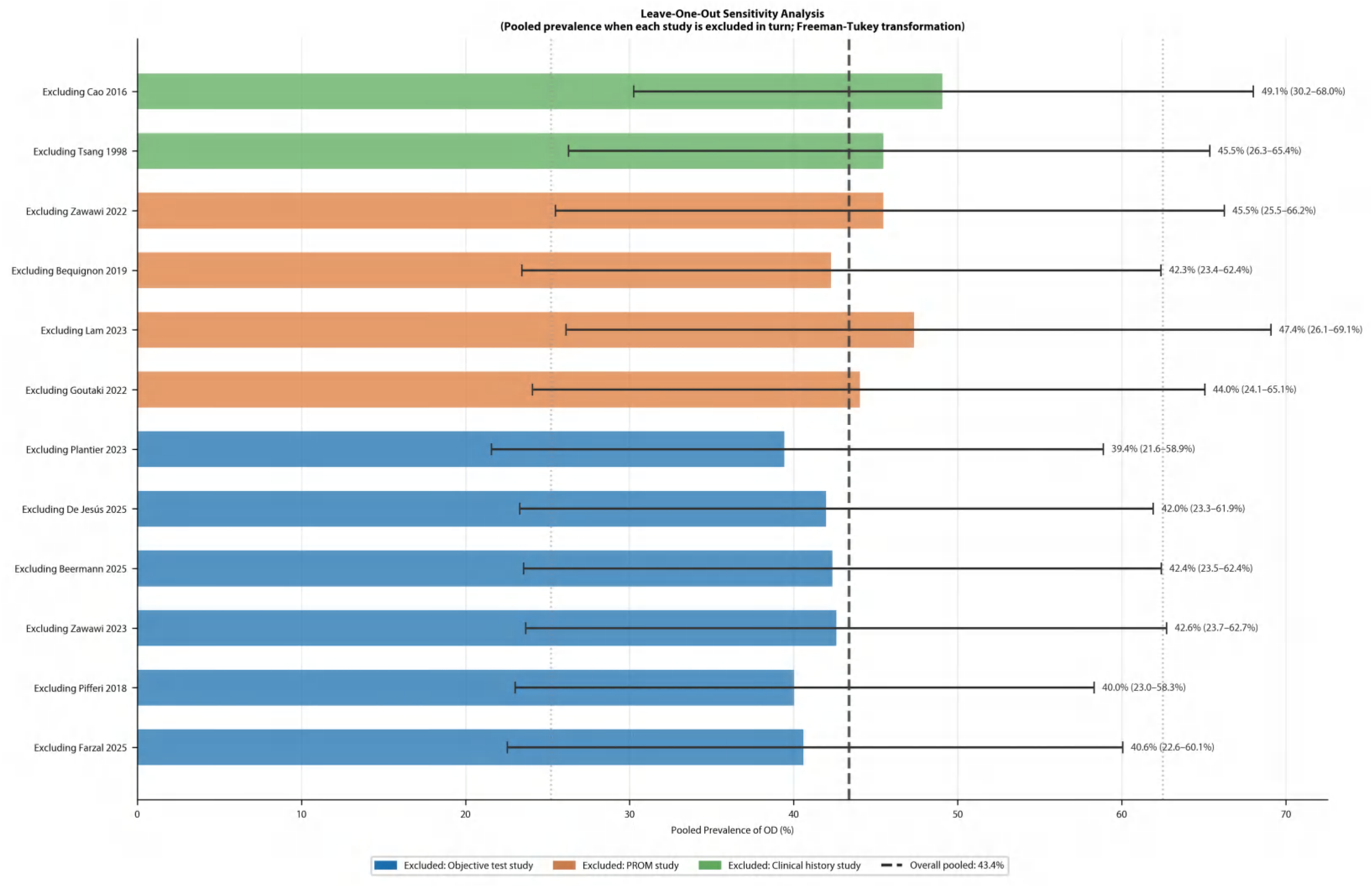
Leave-one-out sensitivity analysis of pooled olfactory dysfunction prevalence in primary ciliary dyskinesia

### Certainty of Evidence (GRADE Assessment)

The certainty of evidence was assessed using the GRADE framework across three outcome domains (Table 2). For the overall pooled prevalence of olfactory dysfunction of 43.4% (95% CI 25.2%–62.5%), the evidence was rated as low certainty, downgraded from high due to serious inconsistency (I²=96.2%, 95% PI 0.1–99.0%) and serious imprecision reflecting the very wide prediction interval. For the objective psychophysical testing subgroup (66.1%, 95% CI 55.5–76.0%), the evidence was rated as moderate certainty, downgraded for risk of bias (two studies rated moderate risk) but not for inconsistency, given the acceptable heterogeneity within this subgroup (I²=40.5%, 95% PI 38.4–88.9%). For the patient-reported outcome measure subgroup (30.5%, 95% CI 11.4–54.0%), the evidence was rated as very low certainty, downgraded for serious inconsistency (I²=95.7%), serious imprecision, and indirectness arising from the use of non-validated and heterogeneous questionnaire instruments across studies.

### Correlates of Olfactory Dysfunction

Several studies investigated factors associated with olfactory function in PCD.

#### Age

Multiple studies reported that olfactory dysfunction in primary ciliary dyskinesia (PCD) worsens progressively with age. This age-related decline is observed in both objective psychophysical testing and patient-reported outcome measures (PROMs). Farzal et al. noted a significant negative correlation between age and UPSIT scores in their cohort of adolescents and adults ^(7)^. Similarly, Goutaki et al. and Lam et al. found that adults reported a higher prevalence of olfactory symptoms than children (48% vs. 17% and 24% vs. 4%, respectively) ^(11,12)^. This progressive decline was also confirmed by De Jesús et al. in their genotype-specific cohort, where age correlated significantly with declining BSIT scores (Pearson r² = 0.29, p = 0.039) ^(13)^. Crucially, De Jesús et al. noted that this age-related decline was not observed in their matched healthy control group, suggesting that the progressive loss is a specific feature of the PCD disease process rather than normal physiological ageing. Regarding sex-specific variations, data are currently limited to a single study. De Jesús et al. reported that females exhibited a much stronger age-related decline in olfactory function (r² = 0.71, p = 0.005) compared to males ^(13)^.

#### Genotype and Ciliary Ultrastructure

Pifferi et al. found that patients with classical TEM abnormalities had worse olfactory scores compared to those with DNAH11 mutations or normal ultrastructure ^(6)^. Farzal et al. observed that individuals with CCDC39 mutations had significantly worse olfactory function than those with CCDC40 mutations^(7)^. Overall, patients with ODA mutations had higher UPSIT scores compared to patients with IDA/MTD and ODA/IDA mutations. In contrast, Zawawi et al. reported normal olfactory function in majority of children with CCDC39 and CCDC40 mutations ^(14)^. No major difference in prevalence of anosmia across mutation types was noted in this study.

#### Sinonasal Disease Burden

Chronic rhinosinusitis was noted in almost all patients ^(15)^. Pifferi et al. demonstrated a significant inverse correlation between the olfactory function described using the TDI scores and the Lund-Mackay score on sinus CT scans, and positive correlation with nNO ^(6)^. For patients with equivalent sinus disease burden, PCD patients had worse olfactory dysfunction compared to non-PCD sinusitis ^(6)^. Beermann et al. reported significant subjective and objective smell impairment in PCD cohort compared to cystic fibrosis patients ^(16)^. Zawawi et al. reported no correlation with median SNOT-22 scores in patients with anosmia and normosmia ^(14)^.

#### Subjective Awareness

A key finding from the studies using objective testing was the high rate of under-recognition of olfactory loss by patients. In the study by Zawawi et al. (2023), 52% of children had objectively measured olfactory dysfunction, yet only 16% of the total cohort had reported any subjective awareness of a smell problem prior to testing^(14)^. Beermann et al. reported a similar, though slightly less pronounced, discrepancy, with 47.4% reporting subjective impairment compared to 55.0% demonstrating objective OD^(16)^.

## Discussion

This systematic review and meta-analysis, the first to our knowledge on this topic, synthesises data from 12 studies (n=865) and confirms that olfactory dysfunction is highly prevalent in primary ciliary dyskinesia (PCD), with an overall pooled estimate of 43.0%. The principal finding is the stark discrepancy between the prevalence of olfactory loss detected by objective psychophysical testing versus subjective reporting. Objective testing reveals a prevalence of 67.3%, whereas patient self-report captures only 30.9%. This discordance between objective reality and subjective perception highlights a significant gap in clinical care and patient awareness, suggesting that a silent majority of individuals with PCD live with an unmanaged sensory deficit.

A large proportion of patients with PCD have concomitant CRS ^(3,15)^. Olfactory dysfunction is common in CRS, with potential impairment of motile cilia in the nasal epithelium as well as non-motile cilia in the specialised olfactory epithelium that is involved in the olfactory signalling cascade ^(17)^. The available evidence points towards a dual pathophysiology driving this sensory deficit, incorporating both a conductive element and a putative primary neurosensory component. The conductive element arises from the physical obstruction of the olfactory cleft by manifestations of PCD-related CRS—viscous mucus, mucosal oedema, and nasal polyposis. Predictably, olfactory dysfunction correlated with extent of sinonasal disease ^(6)^. However, several lines of evidence from this review indicate that the pathophysiology extends beyond simple obstruction. Firstly, Pifferi et al. found that olfactory dysfunction was significantly worse in PCD patients compared to a control group with non-PCD chronic sinusitis, despite a similar radiological burden of sinus disease^(6)^. This crucial finding suggests that the olfactory loss in PCD is intrinsic and not merely a secondary consequence of inflammation. Secondly, the reported associations between major ciliary ultrastructural defects and worse olfactory scores provide a direct link between the fundamental genetic defect and the olfactory phenotype^(6,13)^. There is growing evidence of impairment of olfactory function in other ciliopathies ^(18)^. Higher-powered studies are needed to clarify the mechanistic relationship between ultrastructural defects and the olfactory phenotype, while also controlling for potential confounding effects of CRS.

A consistent finding across multiple studies is the association between increasing age and worsening olfactory function. Age-related decline in olfaction has been established with large normative population studies ^(19)^. De Jesús et al demonstrated an inverse correlation between olfactory function and age using objective testing in PCD population, however, this was not demonstrated in the normal cohort^(13)^. In PCD, as in other forms of CRS, this accelerated age-related decline likely reflects the cumulative lifetime burden of sinonasal inflammation, recurrent infections, and progressive structural damage to the olfactory cleft and neuroepithelium. It is unclear whether olfactory impairment and its subsequent decline with age in PCD patients is solely due to chronic inflammation and mucosal disease, or if certain PCD-related genes are expressed in the olfactory epithelium and uniquely affect olfactory function. Furthermore, age must be considered as a potential confounding factor when interpreting genotype-phenotype correlations. This may explain the normal olfaction noted in the paediatric cohort with CCDC39 mutation in Zawawi et al (2023) study, compared to Farzal et al (2025) study which showed worse olfaction with the same mutation^(7,14)^. Studies comparing olfactory function between different genetic mutations or ultrastructural defects must carefully control for age, as older cohorts will naturally exhibit worse olfactory scores due to cumulative disease burden, potentially masking or exaggerating true genotypic differences.

Another common theme was profound lack of subjective awareness observed in the high-quality studies included in this review. This was well illustrated by Zawawi et al. (2023), in which objective psychophysical testing (U-Sniff) revealed OD in 52% of children, yet only 16% had reported a smell problem subjectively ^(14)^. A companion multicentre study by the same group using the SNOT-22 anosmia item captured only 23.4%, further underscoring the limitations of questionnaire-based screening ^(15)^. Given the congenital nature of PCD, many patients may have experienced olfactory impairment from such an early age that they lack a normal sensory baseline for comparison. In addition, most people are not good at quantifying the strength of their olfactory function^(20)^. This has immediate and serious implications for patient safety, as an unrecognised inability to detect environmental hazards such as smoke, gas leaks, or spoiled food poses a tangible risk. Furthermore, the impact on quality of life—affecting flavour perception, social bonding, and emotional well-being—is significant but may be misattributed to other sinonasal symptoms if not specifically assessed^(21)^.

The substantial heterogeneity (I²=96.2%) across the 12 studies is quantitatively expressed by the very wide 95% prediction interval of 14.4% to 99.8%. While the moderate heterogeneity within the objective testing subgroup (I²=40.5%) suggests that objectively measured olfactory dysfunction is a relatively consistent finding, the very high heterogeneity within the patient-reported outcome measure (PROM) subgroup (I²=95.7%) warrants specific attention. One possible explanation for this wide variance is the use of different, and sometimes unvalidated, questionnaires across studies. This underscores the critical need for future research to employ consistent, validated smell-specific questionnaires to allow for meaningful comparability. For example, the SNOT-22 is validated in both paediatric and adult populations, though it is not exclusively smell-specific. More recently, the Smell-Qx questionnaire has been developed and validated specifically for assessing olfactory and gustatory disorders in adults^(22)^. However, there currently remains a lack of validated smell-specific screening questionnaires designed for paediatric use, representing a significant gap in the assessment toolkit.

### Strengths and Limitations of this Review

The major strength of this review is its comprehensive, PRISMA-compliant methodology, including a prospectively registered protocol and a quantitative meta-analysis that, for the first time, pools the global prevalence of olfactory dysfunction in PCD. The subgroup analysis by assessment method provides a clear, evidence-based explanation for the wide range of prevalence rates reported in the literature.

However, the review has several limitations. The primary limitation is the high degree of statistical heterogeneity (I² = 96.2%) across the included studies. While our subgroup analysis successfully explained a large portion of this by isolating the assessment method as a key variable, significant heterogeneity remained within the PROM and clinical history subgroups. Secondly, the overall number of studies was small (n=12), and many had limited sample sizes, which restricts the statistical power of the meta-analysis and prevented more granular subgroup analyses (e.g., by specific genotype). Thirdly, the cross-sectional design of most included studies allows for the identification of associations but precludes any conclusions about causality or the longitudinal progression of olfactory dysfunction. The fact that several studies were rated as having a moderate or high risk of bias also tempers the strength of the overall conclusions. A further limitation is that data extraction and GRADE assessment were performed by a single reviewer, which introduces the potential for subjective bias.

### Clinical Implications and Future Directions

The results of this review mandate a shift in the clinical management of PCD. Relying on patient-reported symptoms to screen for olfactory loss is demonstrably inadequate. We strongly recommend the integration of routine, objective olfactory screening using validated psychophysical tests (e.g., UPSIT, Sniffin’ Sticks) into the standard multidisciplinary assessment of all individuals with PCD, beginning at diagnosis. A confirmed diagnosis of olfactory dysfunction should trigger specific patient and family counselling regarding safety in the home environment and the potential impact on nutrition and quality of life.

Furthermore, based on the GRADE assessment framework, the certainty of evidence supporting the high prevalence of olfactory dysfunction when measured objectively is moderate, downgraded primarily due to the observational nature of the included studies and the inherent risk of bias. However, the large effect size—the contrast between objective findings and subjective awareness—strengthens the recommendation for routine objective screening.

This review also illuminates a path for future research. There is a pressing need for large, multi-centre, longitudinal studies that follow cohorts of PCD patients from childhood into adulthood, using a standardised battery of objective olfactory tests. Such studies are essential to clarify the natural history of olfactory decline and its relationship with age, genotype, and disease progression. Further investigation into genotype-phenotype correlations, particularly comparing specific ultrastructural defects and their differential impact on olfactory domains while rigorously controlling for age and CRS as confounders, is also warranted. Finally, the development and validation of a smell-specific screening questionnaire for paediatric patients is urgently needed to improve early detection in the clinic.

## Conclusion

Olfactory dysfunction is a highly prevalent yet profoundly under-recognised comorbidity in Primary Ciliary Dyskinesia. The true prevalence is likely masked by an overreliance on subjective reporting, and over two-thirds of those tested objectively may be affected. The pathophysiology is complex, extending beyond simple conductive obstruction to include a primary neurosensory deficit linked to the underlying genetic defect. These findings carry a clear mandate for clinicians to integrate objective olfactory testing into routine PCD care to improve patient safety, quality of life, and our fundamental understanding of this rare disease.

## Data Availability

The data that support the findings of this study are available from the corresponding author upon reasonable request.

## Acknowledgements

Not applicable

## Data sharing and availability

The data that support the findings of this study are available from the corresponding author upon reasonable request. Ethical approval: [Insert Ethical approval statement]

## Author contribution statement

A.Z: data screening, data extraction, data analysis and manuscript preparation; K.L: conceptualization, methodology, data screening, supervision and manuscript review; E.B: conceptualization, methodology, data screening, data extraction, supervision and manuscript review; G.K:, conceptualization, methodology, data screening, supervision and manuscript review. All authors (A.Z, K.L, E.B and G.K) have approved the final version to be published and agree to be accountable for the accuracy and integrity of the contents of this paper.

